# Hanging, strangulation, and suffocation deaths of undetermined intent: Hidden suicides on the rise in post-pandemic Argentina?

**DOI:** 10.1101/2025.10.18.25338282

**Authors:** Carlos M. Leveau

## Abstract

The objective of this study is to identify spatial and spatio-temporal clusters of reported suicides and injury deaths of undetermined intent (DUI) by hanging, strangulation, and suffocation (HAUS); determine whether these clusters coincide geographically; and describe the overlap in their spatiotemporal distributions. Spatial autocorrelation and space-time scan statistics techniques were used to detect spatial and spatiotemporal clusters of HASU-DUI and HASU-suicide in Argentina, during 2015-2023. The spatiotemporal analysis identified distinct geographic clustering of HASU deaths (both suicides and DUI) across Argentina (2015-2023). We observed inverse spatial patterns—low-risk suicide clusters and high-risk DUI clusters expanded—with overlapping areas showing declining suicides and rising DUIs post-2020. Our spatiotemporal analysis suggests potential reclassification patterns between deaths recorded as undetermined intent (DUI) and suicides in post-pandemic Argentina. These findings suggest that combining undetermined deaths by hanging, strangulation and suffocation (Y20) with official suicide data may provide a more complete mortality picture in Argentina.

## Introduction

The Americas represent the only global region where suicide rates have shown a consistent upward trend in recent decades ^1^. Within this context, Argentina maintains one of the highest suicide rates, ranking among the top 10 countries in the region with a stable rate —8 suicides per 100,000 inhabitants— throughout the 2010-2019 period ^2^. After a decline during the COVID-19 pandemic (2020 to 2021), preliminary data suggest Argentina’s suicide rates are returning to pre-pandemic levels ^3^.

However, in Argentina, injury deaths of undetermined intent (DUI) accounted for an estimated 13–17% of all injury-related deaths among women and men, respectively ^4^. The age-adjusted DUI rate showed a substantial increase between 2005 and 2023, rising from 4.6 to 8.4 deaths per 100,000 population ^3^. These patterns suggest that including DUI analysis could provide a more complete understanding of suicide trends in Argentina.

Suicides by hanging, strangulation, and suffocation (HASU) –classified under ICD-10 code X70– represent the predominant mechanism of suicide in Argentina (Figure 1). Deaths of undetermined intent (DUI) involving these mechanisms (ICD-10 code Y20) showed a marked increase from 2015 to 2023, rising from 558 cases in 2015 to a peak of 1,437 in 2022 (Figure 1). This trend contrasts sharply with homicides involving HASU, which remained consistently low (36-61 annual deaths during the same period). The observed temporal patterns suggest that many DUI cases classified as Y20 may in fact represent misclassified suicides.

**Figure 1.**
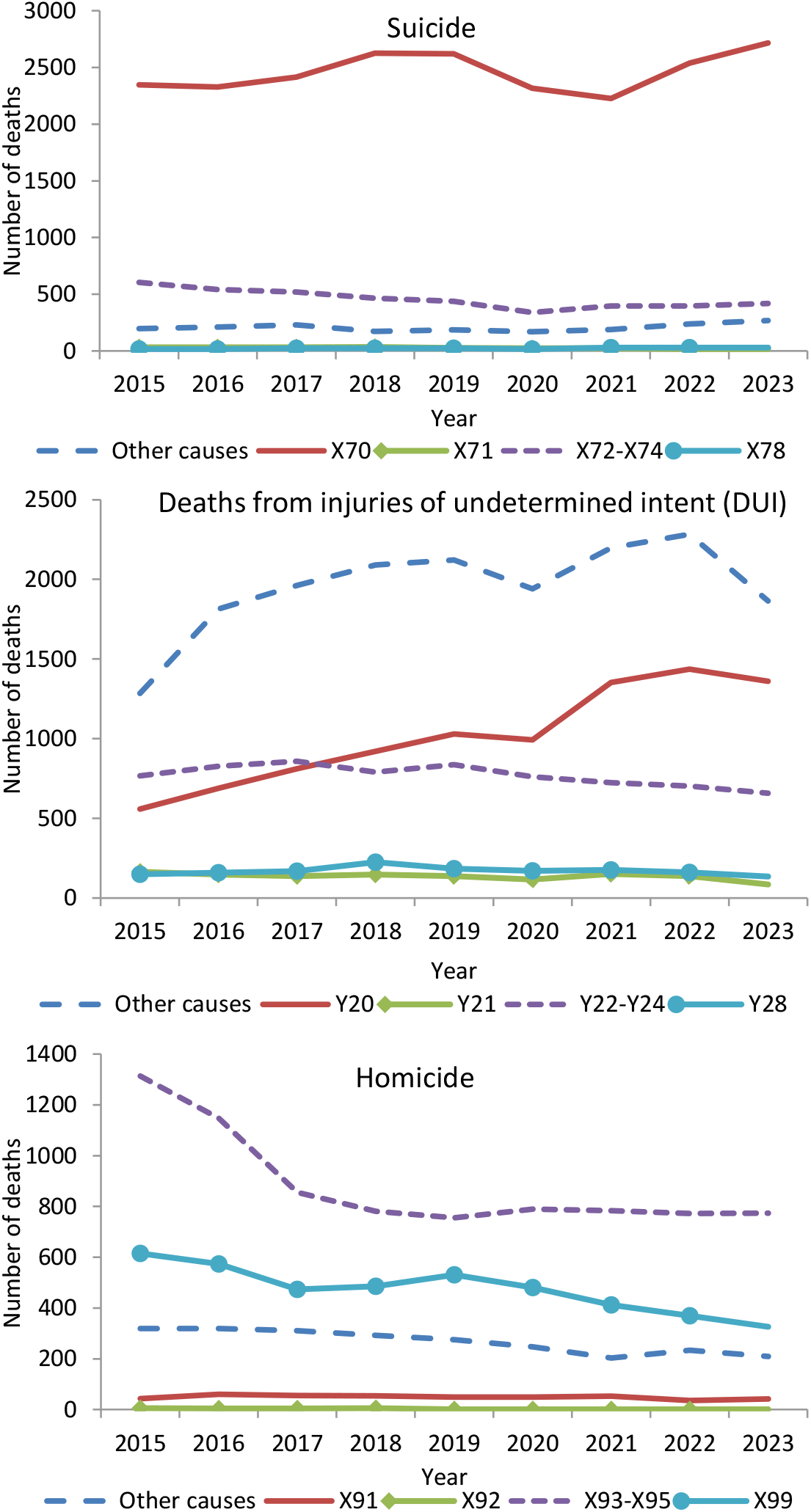
Temporal variation in suicides, deaths from injuries of undetermined intent, and homicides in Argentina, 2015-2023. Note: X70, Y20, X91: hanging, strangulation, and suffocation; X71, Y21, X92: drowning and submersion; X72-X74, Y22-Y24, X93-X95: discharge of firearm; X78, Y28, X99: sharp object; “Other causes” includes other mechanisms of death and deaths from unspecified mechanisms.

While previous research has examined the misclassification of DUI as suicides in Argentina ^4^, no studies to date have specifically investigated suicide underreporting through the lens of HASU –the predominant suicide mechanisms in the country. Existing epidemiological studies on suicide underreporting have primarily adopted temporal approaches ^5–11^, with limited consideration of spatial dimensions ^12–14^. Our study addresses this gap by employing both spatial and spatiotemporal analytical techniques to examine the relationship between officially reported suicides and DUI that share the same mechanism of death classification (HASU).

We propose that suicides and DUI by HASU will show similar spatiotemporal patterns, supporting the hypothesis that many DUIs are misclassified suicides. This study aims to: (1) identify spatial and spatio-temporal clusters of both mortality causes, (2) determine whether these clusters coincide geographically, and (3) describe the overlap in their spatiotemporal distributions. Using spatial quantitative methods, we will compare officially reported suicides (ICD-10 X70) and DUIs (ICD-10 Y20) by HASU in Argentina from 2015 to 2023.

## Methods

We conducted a retrospective ecological study analyzing Argentina’s resident population from 2015 to 2023 (46,234,830 inhabitants recorded in the 2022 national census). We analyzed mortality data from the Directorate of Health Statistics and Information (Ministry of Health) covering suicides (ICD-10: X60-X84), homicides (X85-X99), and DUI (Y10-Y34) at the department level (the smallest administrative division with complete mortality data). In Buenos Aires Province, these units are designated as *partidos*. The study period spanned 2015-2023. Due to incomplete residence data, the Autonomous City of Buenos Aires (CABA) was analyzed as a single spatial unit. The datasets included demographic stratification by sex and age groups for all recorded deaths. We first compared age-stratified mortality frequencies and male-to-female ratios (deaths in men ÷ deaths in women) for suicides, homicides, and deaths of undetermined intent (DUI), focusing on primary death mechanisms. We then performed spatial cluster analysis of hanging, strangulation, and suffocation (HASU) deaths (ICD-10: X70 suicides and Y20 DUI) using spatial empirical Bayes-smoothed annual rates. This empirical Bayes smoothing technique adjusts mortality rates by incorporating spatial dependencies, weighting each department’s observed rate with the general average of neighboring department’s rates ^15^. This approach stabilizes variance estimates, particularly for departments with small populations where random fluctuations may distort mortality rates ^15^. Finally, we analyzed DUI and suicide deaths assuming a Poisson distribution for spatiotemporal analysis. Population denominators, for both spatial and spatiotemporal analysis, were based on: a) 2022 census data, b) linear interpolations between 2010-2022 censuses for 2015-2021, and c) linear projections for 2023.

Moran’s I index was used to measure spatial autocorrelation (geographic clustering) in suicide and DUI risk at the department/*partido* level ^15^. The index ranges from −1 to 1, where positive values indicate clustering of similar values (high-risk or low-risk areas), negative values suggest dispersion (dissimilar values cluster), and values near zero imply random spatial distribution. Adjacency between geographic units was defined using Queen contiguity, which considers units sharing boundaries or corners as neighbors. To assess spatial autocorrelation of suicide/DUI risk due to HASU, we performed permutation tests (n=9999) to generate random distributions of Moran’s I under the null hypothesis of spatial independence. The observed Moran’s I was considered statistically significant (p<0.05). While Moran’s I measures global spatial autocorrelation, it does not identify specific cluster locations. We therefore calculated Local Indicators of Spatial Association (LISA) to detect four types of spatial clusters ^15^: i) departments with elevated suicide/DUI risk surrounded by neighboring departments with similarly high risk (high-high); ii) high-risk departments adjacent to low-risk areas (high-low); iii) low-risk departments surrounded by high-risk neighbors (low-high); and iv) departments with low risk bordering other low-risk areas (low-low). We analyzed spatial co-occurrence between DUI and suicide mortality by HASU using bivariate LISA clustering. This identified four patterns: i) high HASU-DUI with high HASU-suicide neighbors (“high-high”); ii) high HASU-DUI with low HASU-suicide neighbors (“high-low”); iii) low HASU-DUI with high HASU-suicide neighbors (“low-high”); and iv) low rates of both (“low-low”). High-high clusters, for example, represented departments with elevated HASU-DUI rates surrounded by departments with high HASU-suicide rates.

We used space-time Poisson models ^16^ to detect spatiotemporal clusters of suicide and DUI deaths in Argentina (2015-2023). This approach identifies areas with significantly higher or lower mortality risk during specific time periods compared to the study area. The analysis incorporated both temporal (year of death) and spatial (department of residence) dimensions. Using scan statistics, we compared observed versus expected deaths within variable temporal windows and adjustable circular geographic areas across all possible department-year combinations ^16^. We established maximum spatial and temporal scanning thresholds at 10% of the population and 50% of the study period, respectively. A 10% population threshold was selected to optimize detection of localized clusters. Clusters were identified as cylinders with geographic (base) and temporal (height) dimensions. Each cluster’s relative risk (RR) indicated excess mortality (e.g., RR = 1.50 = 50% more suicides in that cluster compared to the rest of Argentina during the same period). Statistical significance (p < 0.05) was determined using Monte Carlo tests with 9,999 permutations ^16^.

All spatial and spatiotemporal analyses were performed using GeoDa (Center for Geospatial Analysis and Computing, Arizona State University) for Moran’s I and LISA statistics, SaTScan v9.4.4 for spatiotemporal clustering ^16^, and QGIS 2.14.3 for spatial visualization of results.

## Results

From 2015 to 2023, Argentina’s national mortality records documented 15,118 homicide deaths, 36,385 deaths from injuries of undetermined intent (DUI), and 28,590 suicides. Analysis of death mechanisms revealed distinct patterns across these categories (Figure 2). Suicides were predominantly caused by HASU, though the proportion of HASU deaths decreased with advancing age. In contrast, firearm injuries constituted the most frequent homicide mechanism in all age groups except those aged ≥65 years, with decreasing prevalence among older adults. Among DUI, both firearm-related deaths and HASU showed inverse relationships with age, suggesting these may represent misclassified homicides and suicides, respectively (Figure 2). Male-to-female mortality ratios showed closest correspondence between suicides and DUI by HASU, suggesting potential misclassification between these categories (Figure 3). Conversely, firearm- and sharp object-related deaths demonstrated similar sex ratios between homicides and DUI, indicating these DUI cases may represent unreported homicides (Figure 3).

**Figure 2.**
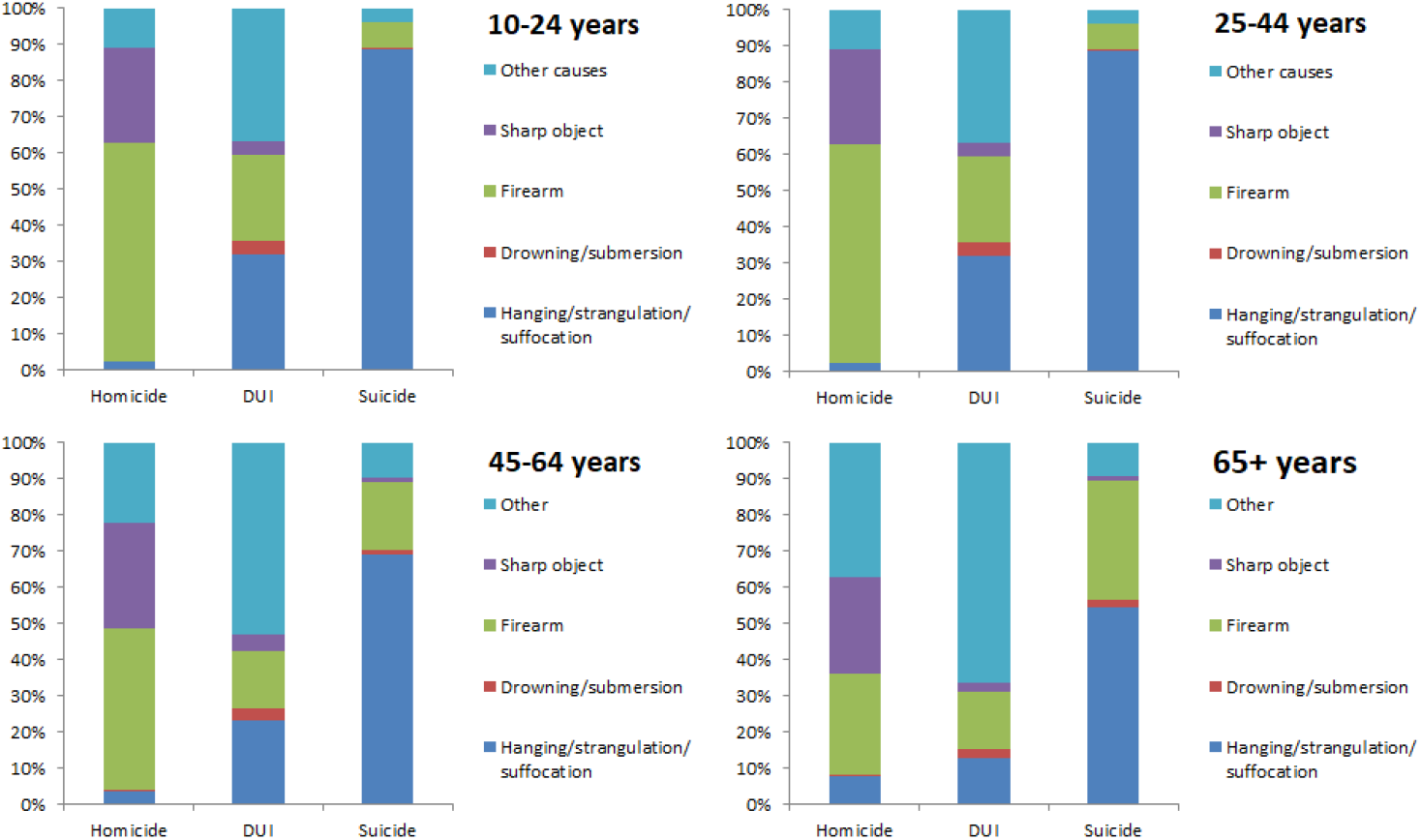
Percentage distribution of the main mechanisms of death in homicide, DUI, and suicide. Argentina, 2015–2023.

**Figure 3.**
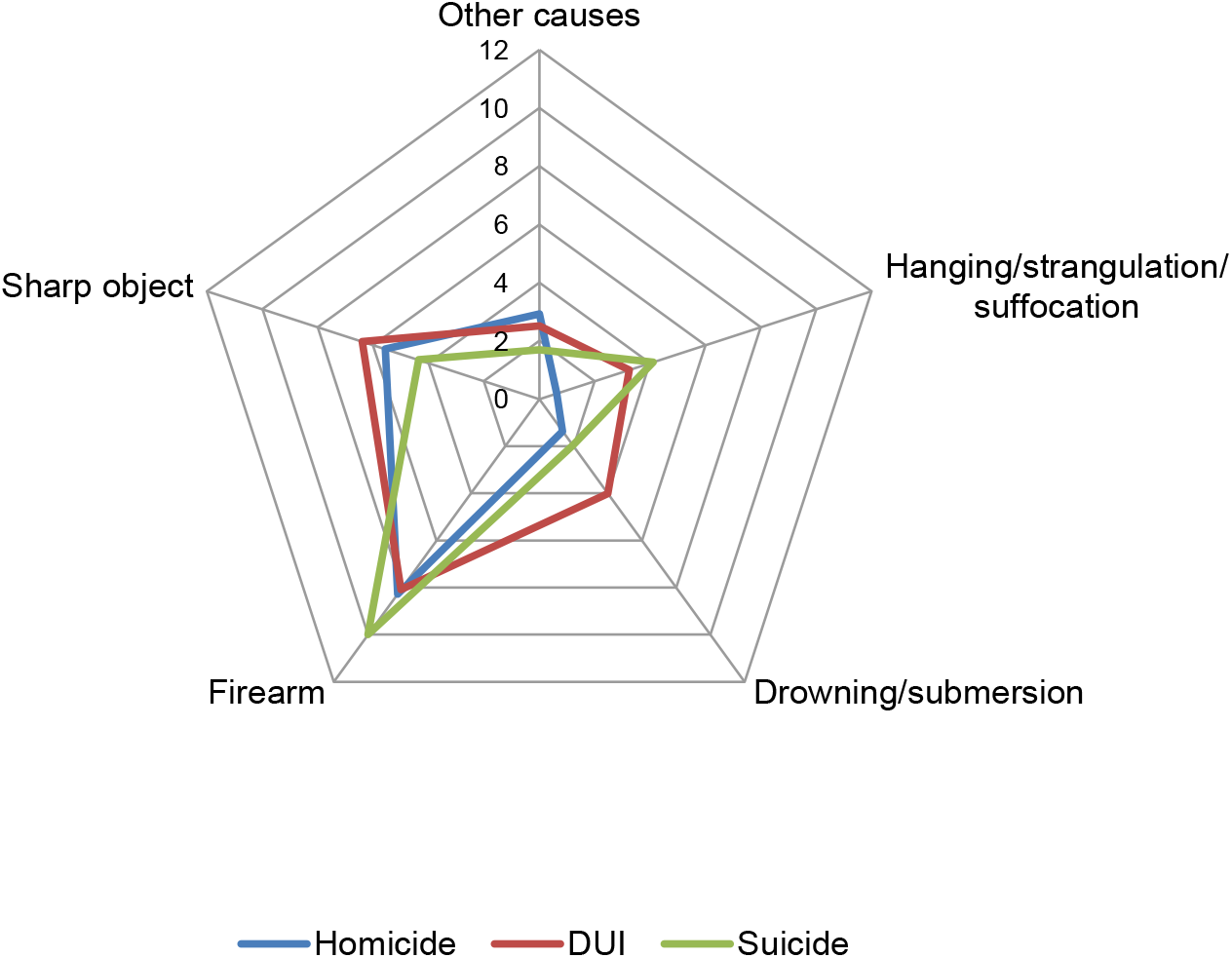
Sex ratios—the ratio of male deaths to female deaths—of the main mechanisms of death in homicide, DUI, and suicide. Argentina, 2015–2023.

Figure 4 shows the spatial clusters of HASU-suicide in Argentina. The largest geographic clusters of high suicide risk were persistently located in the northwest of the country, clusters occurring intermittently in southern Patagonian provinces (Chubut, Neuquén, and Santa Cruz). Low-suicide-risk clusters were concentrated in Buenos Aires, Córdoba, and southern Santa Fe provinces. From 2020-2023, the Greater Buenos Aires cluster expanded to cover eastern Buenos Aires province (Figure 4).

**Figure 4.**
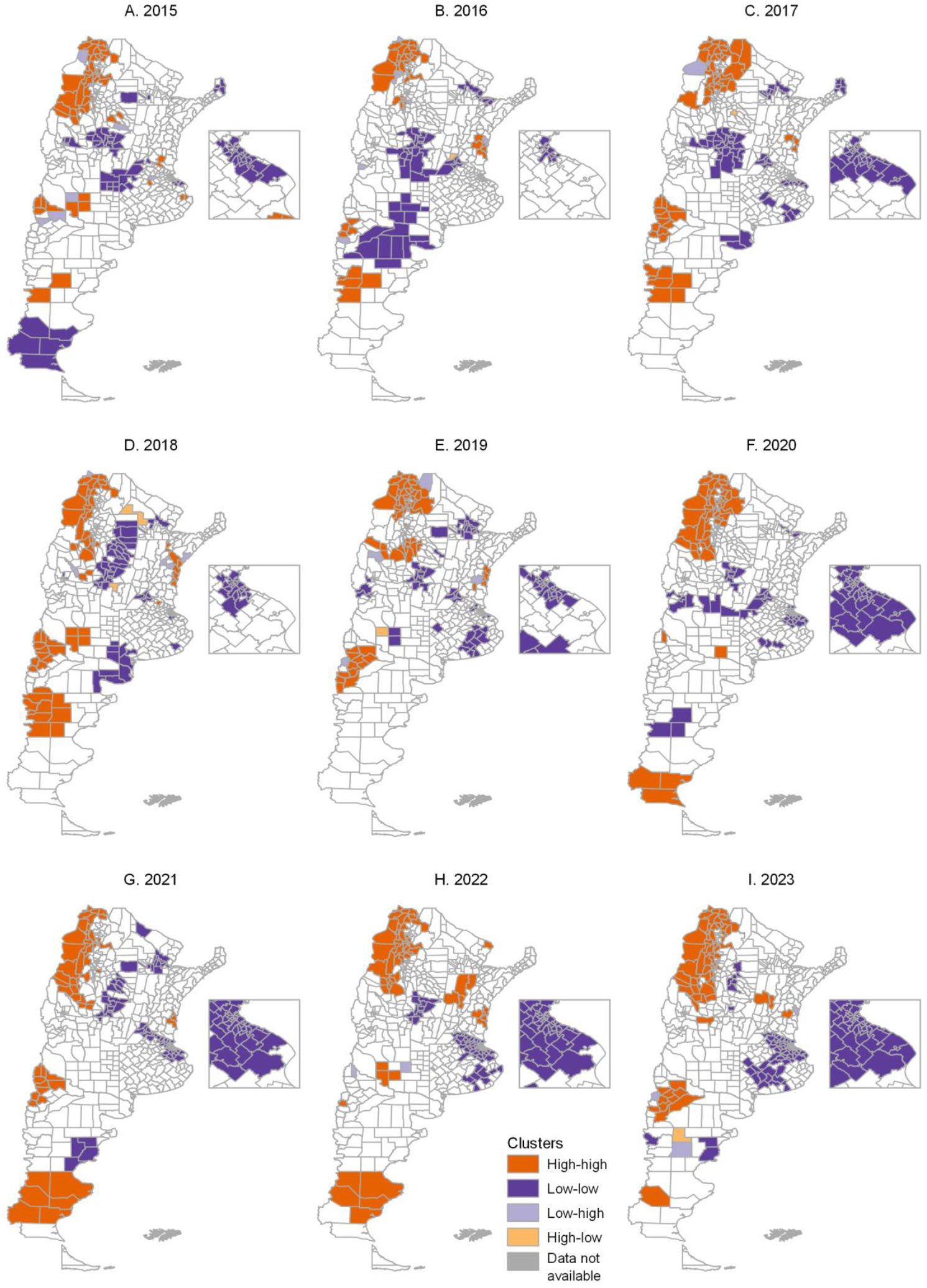
Spatial clusters of HASU-suicide. Argentina, 2015–2023.

Persistent high HASU-DUI risk clusters occurred in Buenos Aires, Córdoba, and southern Santa Fe (Figure 5) -regions with low HASU-suicide risk (Figures 4). Furthermore, Chaco province showed concurrent high DUI risk (2015-2021) and low suicide risk (2016-2021), suggesting potential misclassification (figures 4-5). For a more detailed description of the geography of mortality risk from HASU-suicides and HASU-DUI, Figures S1 and S2 (Supplementary Material) show the smoothed annual crude rates, respectively.

**Figure 5.**
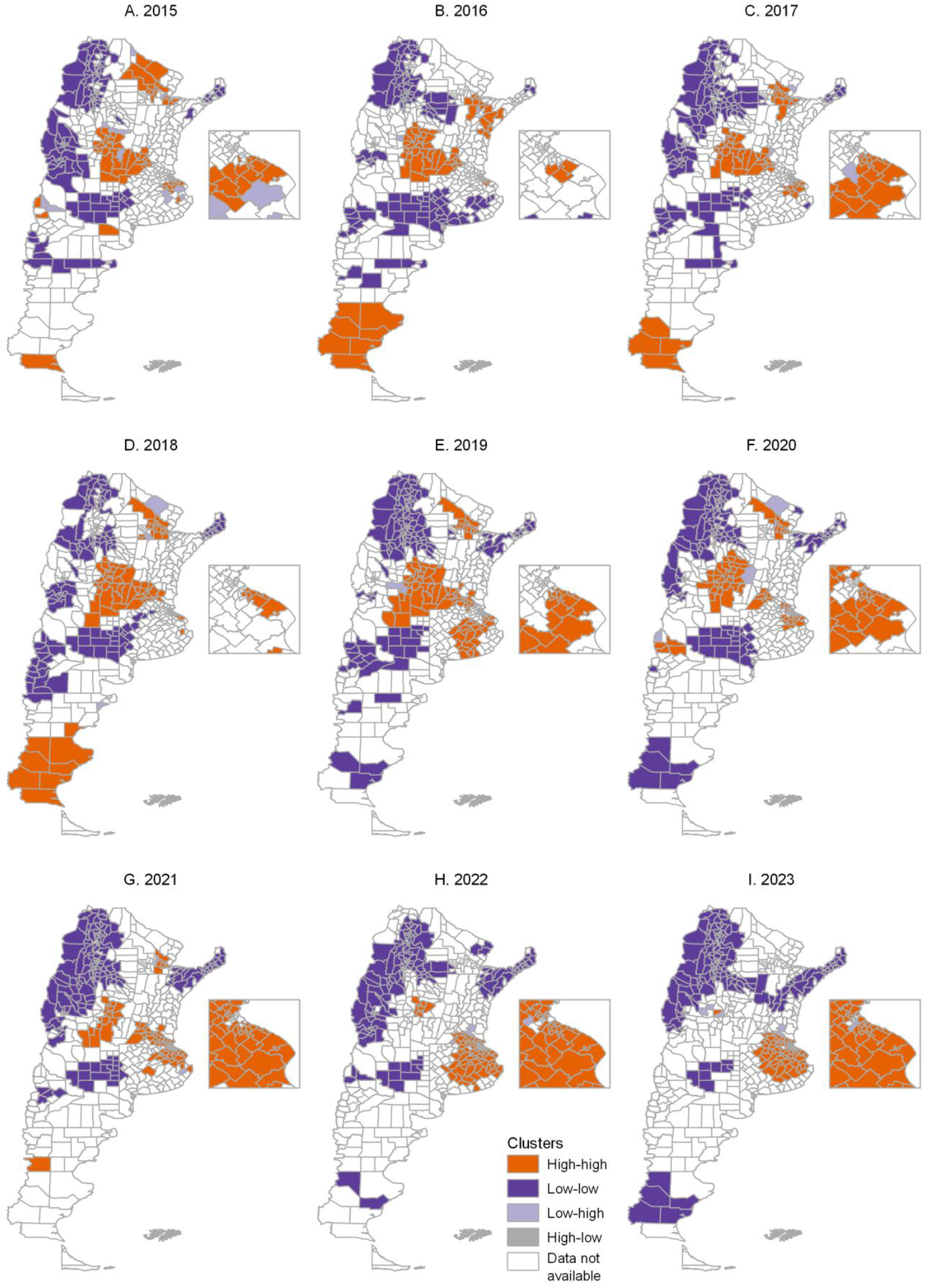
Spatial clusters of HASU-DUI. Argentina, 2015–2023.

The bivariate Moran’s I analysis revealed peak spatial autocorrelation between HASU-DUI and HASU-suicide during 2021-2023 (Figure 6). This reflected two concurrent patterns: (1) the geographic diffusion of HASU-DUI within Buenos Aires Province, and (2) this diffusion occurring specifically in areas with low HASU-suicide risk (Figure 6).

**Figure 6.**
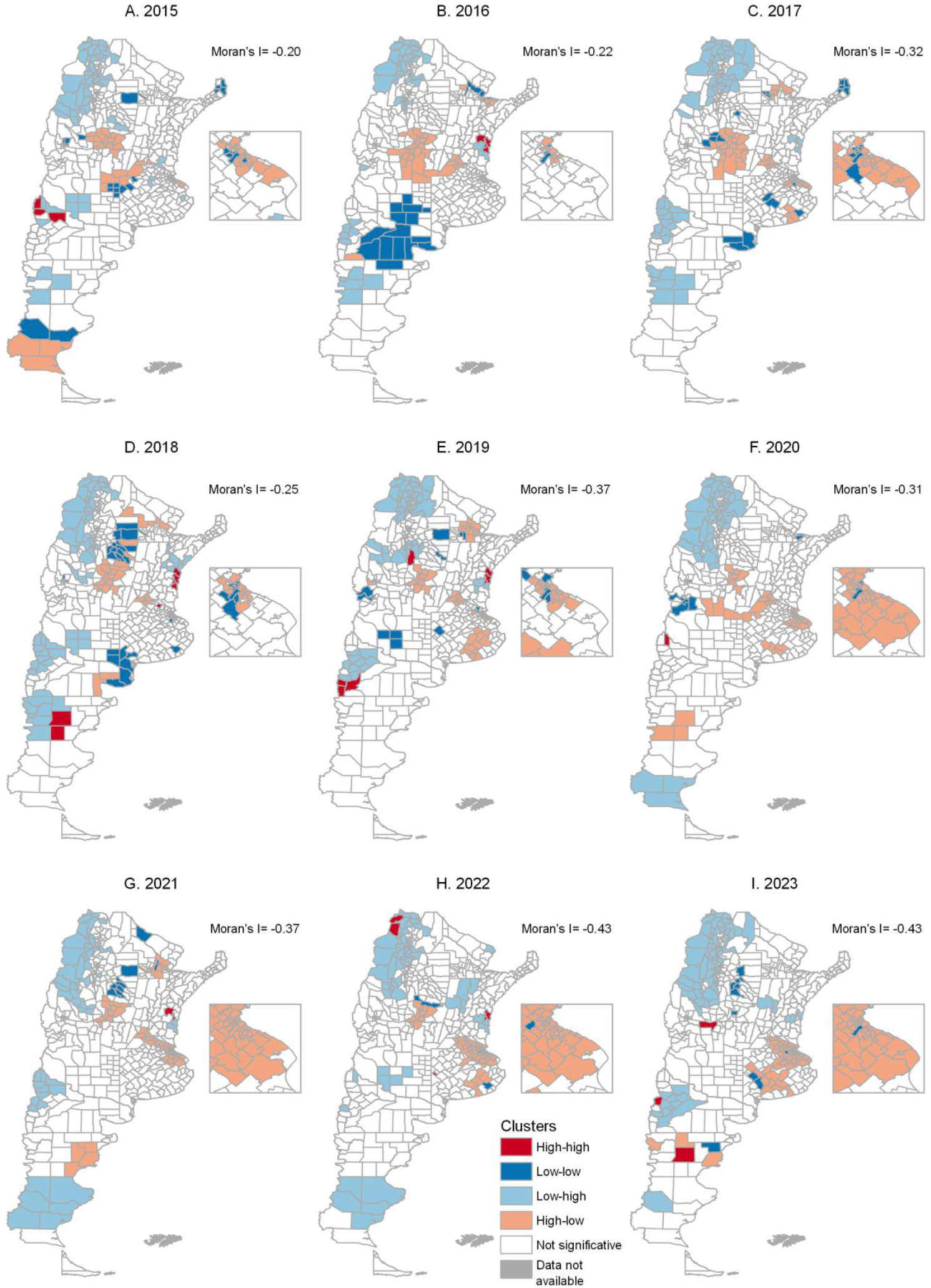
Bivariate spatial clusters between HASU-DUI and HASU-suicide. Argentina, 2015– 2023.

Spatiotemporal analysis revealed that 70% of suicide clusters (7/10) emerged during pandemic and post-pandemic years (2020–2023) (Figure 7.A). A similar proportion was observed for DUI clusters, though with an inverse geographic distribution: areas with high (or low) suicide risk consistently corresponded to areas with low (or high) DUI risk (Figure 7.B). Figure 7.C shows the geographic overlap between clusters of both causes of death. In several of these areas of geographic overlap in central Argentina, a coincidence is observed between clusters showing a decrease in suicides during the pandemic and post-pandemic periods, and clusters showing an increase in undetermined deaths during the same period. These overlapping areas show an inverse temporal pattern: locations with declining suicide risk during and post-pandemic (2020– 2023) concurrently exhibited increasing DUI risk. This reciprocal relationship suggests possible HASU-suicide misclassification during the study period.

**Figure 7.**
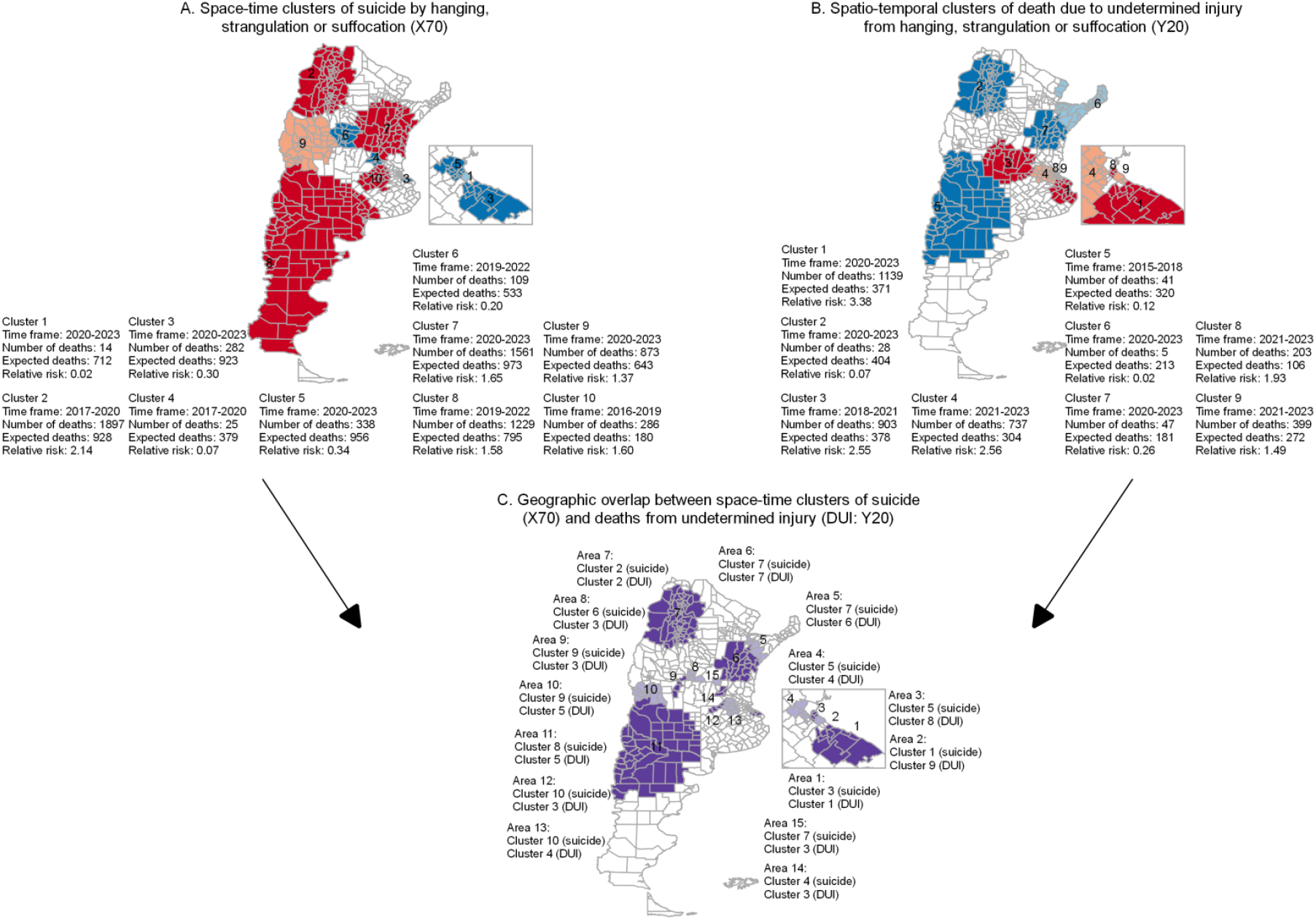
Spatiotemporal clusters of HASU-DUI, HASU-suicide, and areas of geographic overlap between the two cluster types. Argentina, 2015–2023.

Figure 8 displays mortality trends in overlapping HASU-suicide and HASU-DUI clusters, revealing two distinct patterns across geographic areas: sustained high versus low mortality rates persisting for ≥2 years. In six areas (75%), inverse trends emerged: suicide decreases corresponded with DUI increases, and vice versa. Three areas (1, 3, 4) showed significant DUI rises beginning 2019-2020 alongside sustained suicide declines through 2023. The inverse relationship was most pronounced in Area 8 during 2023, where a sharp DUI decline mirrored an equivalent suicide increase. When analyzing combined suicide and DUI deaths by HASU, six areas (1, 2, 3, 4, 8, and 14) demonstrated distinct temporal patterns compared to suicide alone (Figure 8). Areas 1, 3, and 4 exhibited increasing HASU deaths (suicides + DUI) alongside decreasing suicide-only deaths during the same period.

**Figure 8.**
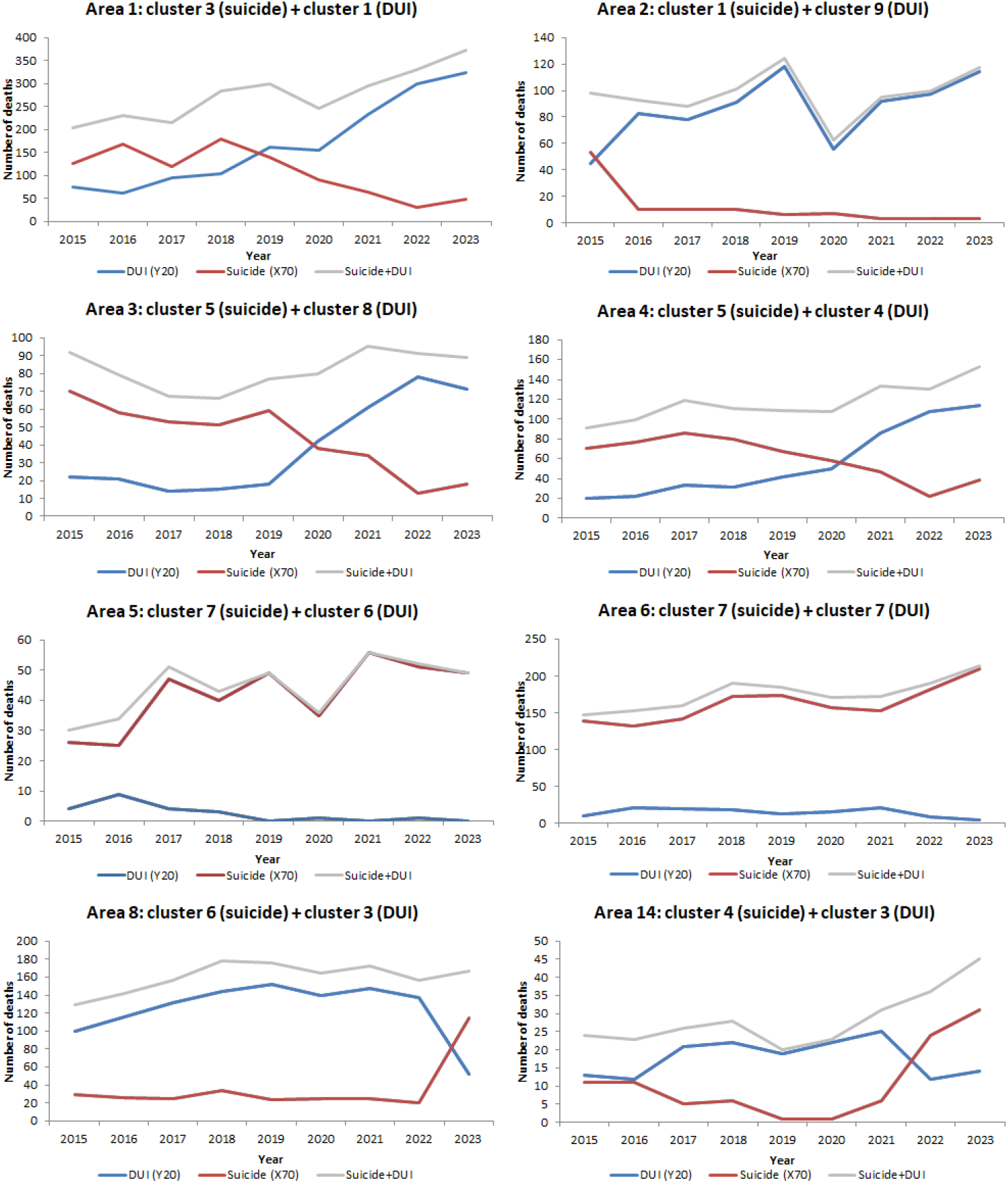
Number of HASU-suicides (X70) and HASU-DUIs (Y20) per year in areas where spatiotemporal clusters of both causes of death overlap. Argentina, 2015–2023. See Figure 7.C for the location of overlap areas.

## Discussion

From 2015–2023, suicide deaths in Argentina were predominantly caused by hanging/strangulation/suffocation (HASU), while homicides were primarily firearm-related. Both mechanisms remained the leading causes in their respective categories despite decreased prevalence among those aged ≥65 years. Sex-ratios patterns revealed significantly closer correspondence between HASU deaths classified as DUI versus suicides compared to homicides. This similarity in sex distribution between HASU-DUI and HASU-suicide suggests potential misclassification bias in mortality coding. Our spatiotemporal analysis identified distinct geographic clustering of HASU deaths (both suicides and DUI) across Argentina (2015-2023). We observed inverse spatial patterns—suicide clusters contracted while DUI clusters expanded—with overlapping areas showing declining suicides and rising DUIs post-2020. This study provides the first national-level comparison of these mortality distributions employing spatial epidemiological techniques.

Argentina’s 2023 suicide rate ^3^ (7.5/100,000) increased 39% (to 10.3/100,000) when including hanging/strangulation/suffocation (HASU) deaths of undetermined intent. While official rates remained stable (7.4-7.5; 2015-2023) with a pandemic dip ^3^, suicide rates including HASU-DUI showed a steady rise from 8.7 (2015) to 10.3 (2023), interrupted only during COVID-19 (2020-2021). The underreporting effect was particularly pronounced in areas with overlapping suicide and DUI clusters. Area 1 exemplifies this pattern: while reported HASU suicides decreased by 83% (from 178 in 2018 to 30 in 2022), the combined HASU mortality (suicides + DUI) increased by 83% (203 in 2015 to 372 in 2023). This divergence reveals substantial misclassification concentrated in specific geographic-temporal clusters.

Determining the cause of violent deaths remains challenging without definitive evidence (e.g., suicide notes), justifying the use of undetermined intent (DUI) classifications ^17^. However, our analysis reveals non-random geographic patterns where declining rates of HASU-suicides precisely coincide with rising HASU-DUI rates in the same areas and time periods. The rapid increase and geographic diffusion of hanging/strangulation/suffocation (HASU) deaths classified as undetermined intent is particularly notable, as these violent mechanisms typically exhibit lower misclassification rates than more ambiguous methods like poisoning ^18^. The spatial and spatiotemporal analyses reveal patterns suggesting non-random geographic diffusion in death certification practices, particularly the systematic underreporting of suicide intent. This may suggest physicians’ concerns regarding statistical confidentiality protocols and perceived professional risks when documenting suicides ^19^.

The bivariate spatial analysis revealed peak spatial autocorrelation (2022–2023) between HASU-DUI rates and HASU-suicide rates, with a prominent “high-low” cluster (high HASU-DUI adjacent to low HASU-suicide rates) encompassing much of Buenos Aires Province and CABA (Figure 6). Spatiotemporal analysis further identified overlapping clusters where HASU-suicide rates declined and HASU-DUI rates increased proportionally, indicating geographic diffusion of HASU-DUI that occurred primarily in eastern Buenos Aires (2021–2023). These synchronized inverse trends suggest systematic reclassification of HASU-related deaths from suicide to DUI in these regions during the study period. The geographic spread of HASU-DUI appears to have originated in urban municipalities of southern Greater Buenos Aires and Greater La Plata before expanding to less densely populated areas of Buenos Aires province. Despite recent improvements in vital statistics quality, this region continues to report a high proportion of deaths classified under “unusable causes of death” ^20^—a category that includes injuries of undetermined intent and provides minimal etiological information.

The hypothesis that suicide rates would rise following the COVID-19 pandemic emerged early, driven by anticipated psychosocial consequences—including fear of contagion, economic instability, and prolonged social isolation ^21^. Our analysis revealed spatial contraction in 2020 for undetermined deaths (DUI) by hanging, strangulation or suffocation within both the Buenos Aires and Córdoba clusters. This synchronized decline in DUI alongside confirmed suicides suggests an authentic reduction in suicide mortality during Argentina’s strict lockdown period, rather than classification artifacts.

This study has several limitations. First, our analysis was restricted to deaths by hanging, strangulation, or suffocation (classified under X70 for suicide and Y20 for undetermined intent), which excludes other important mechanisms of violent death. Given that poisoning (X60-X69) accounts for 4% of all suicides (11% among women) in Uruguay ^22^—a country with sociodemographic and cultural similarities to Argentina—underreporting of suicide by this mechanism in Argentina appears likely, particularly in female populations.

Second, not all hanging deaths are suicides, and among strangulation deaths coded as undetermined intent (Y20), a substantial proportion may reflect homicides.

Third, while disaggregated analysis by sex and age groups was precluded by data limitations (department-level spatial units would introduce excessive zero-inflation), our findings nonetheless demonstrate clear evidence of suicide-to-DUI reclassification in areas exhibiting significant temporal mortality shifts.

Our spatiotemporal analysis suggests potential reclassification patterns between deaths recorded as undetermined intent (DUI) and suicides in post-pandemic Argentina. These observed trends may have implications for suicide surveillance, particularly given that hanging/strangulation/suffocation (HASU) represented approximately three-quarters (77%) of recorded suicides during 2015–2023. While these patterns are consistent with possible misclassification, further validation would be needed to confirm their interpretation. Our findings suggest that combining undetermined deaths by hanging, strangulation and suffocation (Y20) with official suicide data may provide a more complete mortality picture in Argentina. To improve cause-of-death accuracy, potential strategies could include enhanced physician training programs and systematic cross-verification with complementary data sources like medical records ^20^.

## Data Availability

All data produced are available online at https://www.datos.gob.ar/dataset

## Statements and Declarations

## Consent for publication

Not applicable.

## Ethics approval and consent to participate

Not applicable.

## Supplementary Material

**Figure S1.**
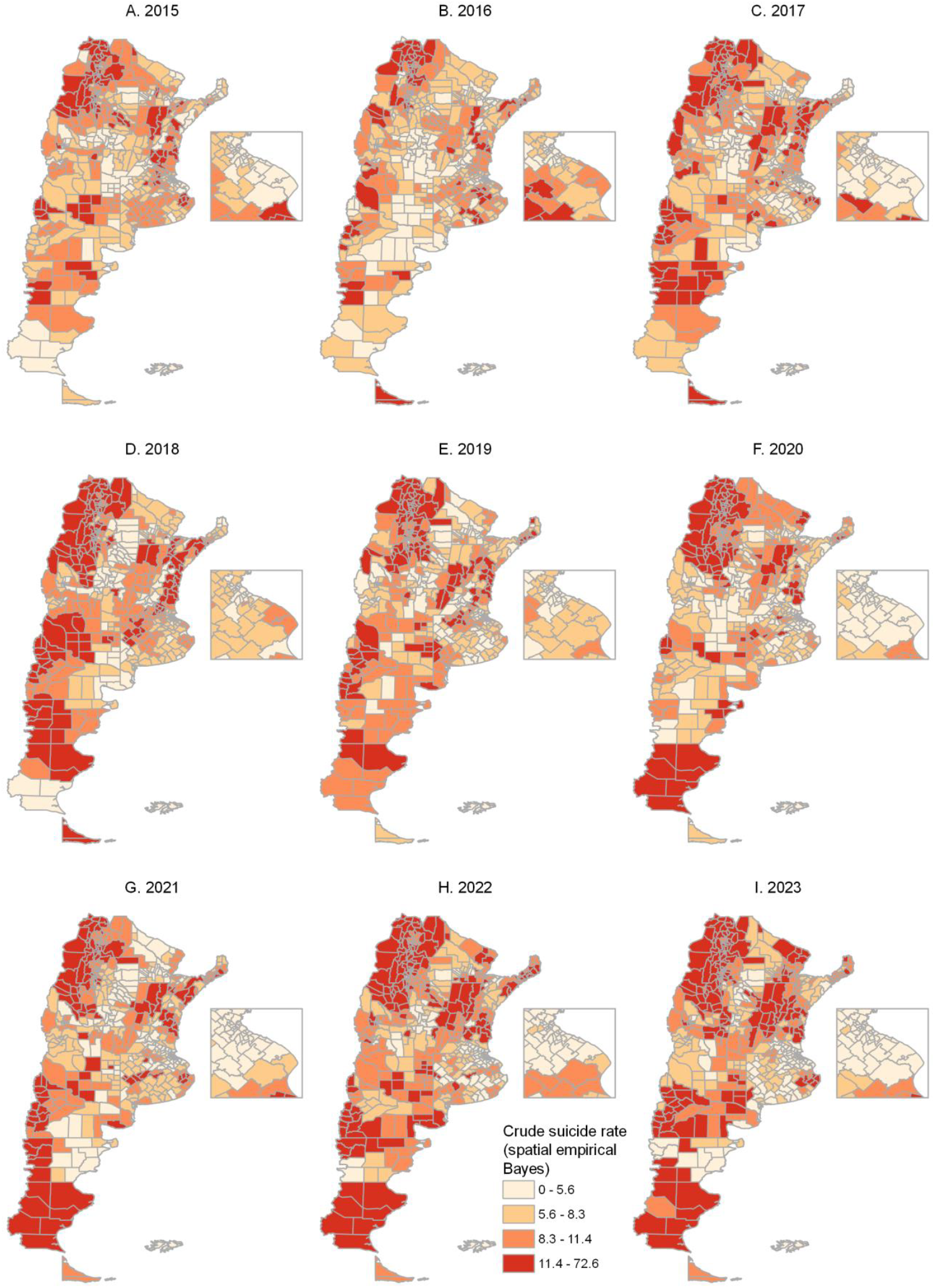
Smoothed* HASU-suicide (X70) rates (deaths per 100,000 inhabitants). Argentina, 2015– 2023. *Smoothed rates using the spatial empirical Bayes technique.

**Figure S2.**
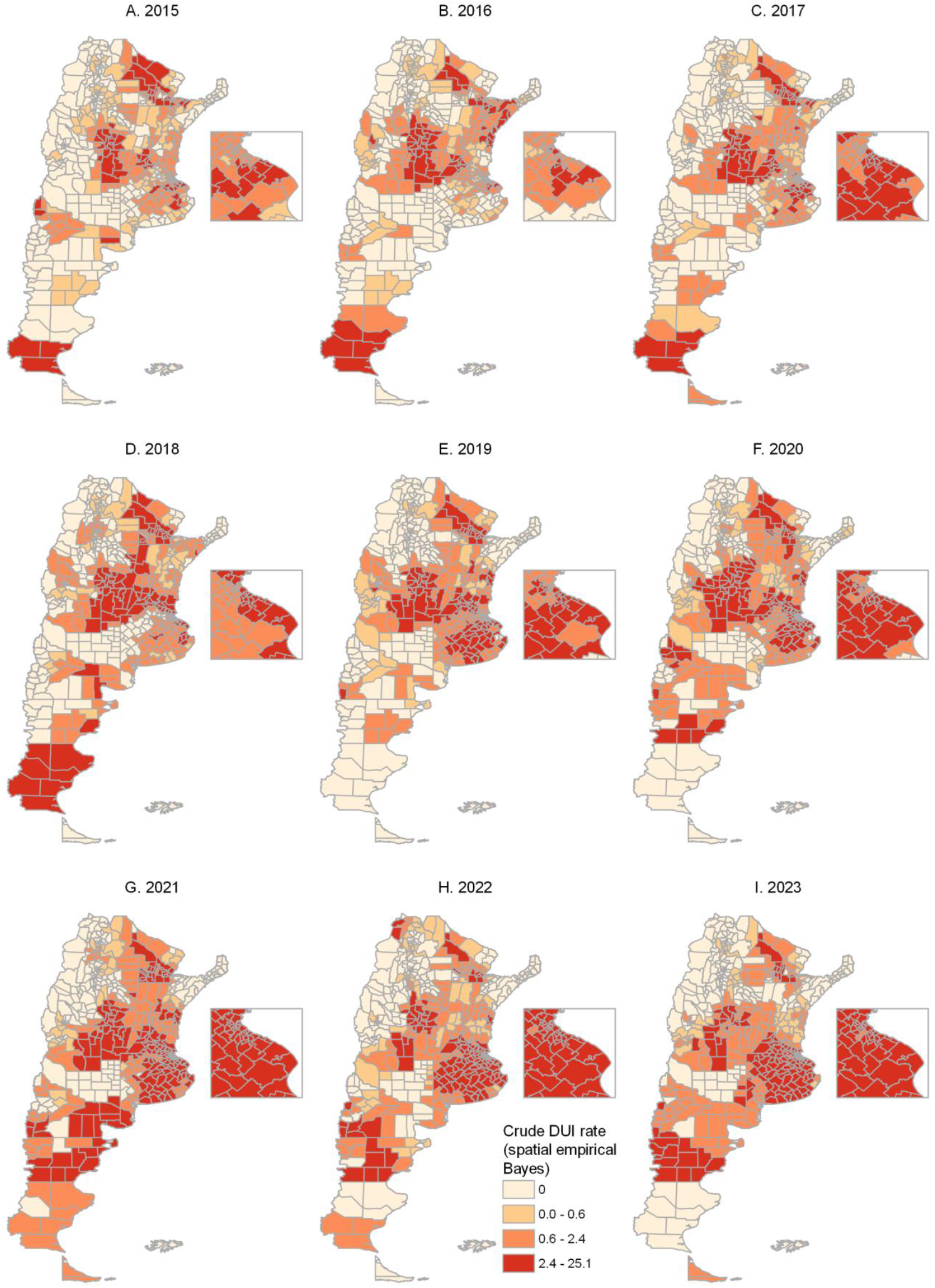
Smoothed* HASU-DUI (Y70) rates (deaths per 100,000 inhabitants). Argentina, 2015–2023. *Smoothed rates using the spatial empirical Bayes technique.

